# Sickle cell disease as a multisystem risk factor: comparative rates of 30 acute and chronic conditions in the United States

**DOI:** 10.64898/2026.11.23.26368722

**Authors:** Taylor S. Noyes, Ni Gusti Ayu Nanditha, Corey Teply, Theresa A. McHugh, Ksenia I Linville, Ihunanaya Okorie, Jack T Fox, Ashley A. Harris, Catherine Chen, Kelly Cercy, Jackie Markt-Maloney, Aishe Memetova, Heather Taylor, Nandita Perumal, Jonathan F. Mosser, Christopher J.L. Murray, Simon I. Hay, Bosede B. Afolabi, Zulfiqar A. Bhutta, Domenico Girelli, Khaled M. Musallam, Frédéric B. Piel, Emily R. Smith, Parminder S. Suchdev, Léon Tshilolo, Nicholas J. Kassebaum

**Affiliations:** Institute for Health Metrics and Evaluation, University of Washington, Seattle, WA, USA; Department of Anesthesiology and Pain Medicine, University of Washington, Seattle, WA, USA; Department of Epidemiology and Biostatistics, University of South Carolina, Columbia, SC, USA; Department of Health Metrics Sciences, University of Washington, Seattle, WA, USA; Pediatricc Infectious Diseases, Seattle Children’s Hospital, Seattle, WA, USA; Center for Global Child Health, Hospital for Sick Children, Toronto, Canada; Center of Excellence in Women and Child Health, Aga Khan University, Karachi, Pakistan; University of Lagos, Department of Obstetrics and Gynaecology, College of Medicine, PMB 12003, Idi-Araba, Lagos, Nigeria; Section of Internal Medicine, Department of Medicine, University of Verona, Policlinico Giambattista Rossi, Verona, Italy; Center for Research on Rare Blood Disorders (CR-RBD), Burjeel Medical City, Abu Dhabi, United Arab Emirates; Department of Global Health, Milken Institute School of Public Health The George Washington University, Washington, DC, USA; Emory Global Health Institute, Emory University, Atlanta, GA, USA; Center for Global Health, US Centers for Disease Control and Prevention, Atlanta, Georgia, USA; Centre Hospitalier Mère-Enfant Monkole, Kinshasa, Democratic Republic of the Congo; Centre de Formation et d’Appui Sanitaire (CEFA), Mont-Ngafula, Kinshasa BP 817, Democratic Republic of the Congo; Imperial College London, Epidemiology & Biostatistics, London, United Kingdom; Department of Global Health, University of Washington, Seattle, WA, USA

## Abstract

Sickle cell disease (SCD) is associated with widespread multi-organ comorbidity, but these outcomes have never been quantified comprehensively against a population without hemoglobinopathies or hemolytic anemias. That gap limits clinical appreciation of the breadth of SCD-associated illness and, because burden-of-disease frameworks require exposure-outcome effect sizes, it also leaves SCD systematically under-counted in global health estimates. Using longitudinal utilization and demographic data from MarketScan for 2016 to 2019, we fitted Bayesian meta-regressions of comparative rates of 30 acute and chronic diseases among individuals with and without an SCD diagnosis, by age and sex. Individuals with SCD had elevated risk of nearly every outcome, with the highest relative risk (RR) and incidence rate ratio (IRR; per person-year) generally in the youngest age groups. This included well-established problems such as chronic pain (RR=12.0, 95% UI 11.0-13.0), but also conditions in which impaired perfusion or persistent vascular injury may be causal: preeclampsia (IRR=9.7, 7.8-12.1), postpartum hemorrhage (IRR=13.6, 11.2-16.1), pulmonary hypertension (RR=96.3, 85.9-108.1), acute myocardial infarction (IRR=28.7, 24.5-34.3), ischemic stroke (RR=16.7, 15.0-18.6), kidney disease (RR=11.9, 10.9-13.0), asthma (RR=3.21, 2.98-3.44) and lower respiratory infections (IRR=2.85, 2.69-3.02). These results supply the inputs needed to quantify indirect SCD burden in comparative risk assessment frameworks. These results establish SCD as a whole-body risk factor rather than a single-organ disease, which should inform treatment guidelines and population-level health programs in settings where SCD is endemic.

## Introduction

Sickle cell disease (SCD) refers to an autosomal recessive disorder of hemoglobin that affects millions of people worldwide.[1–3] Depending on genetic, socioeconomic, and environmental factors, people born with SCD experience subsequent health complications, ranging from more frequent acute events to chronic complications that can lead to multi-organ failure and early death.[2,4,5] Over 1,000 babies are born with SCD in the US annually, where increasing life expectancy of those with SCD has led to steady increases in the size of the SCD population between 2000 and 2021.[6]

Life expectancy of people with SCD has increased substantially in recent decades due to advances in medical knowledge and technology, alongside novel therapies. In the US, life expectancy of individuals with SCD increased from 20 years of age in 1970 to 52 years between 2008 and 2016.[7,8] Nevertheless, mortality among pregnant women with SCD remains 10–26 times greater than in pregnant women without SCD, a statistic which has not improved, even when compared to decades-old data.[9–11] Hydroxyurea remains the most widely available and primary treatment for SCD.[12–14] Transformative gene therapy treatments are available, though likely out of reach for the vast majority of patients—with US list prices of approximately $2 million per treatment.[15] Consequently, there is a heightened prevalence of chronic comorbidities within this population, underscoring the need to understand the magnitude of increased disease risk.

Sickle cell hemoglobin (HbS) polymerization causes malformed red blood cells and is a defining feature of SCD. The sickling of red blood cells causes obstructed blood flow, hemolysis, and other cellular abnormalities that reduce oxygen delivery to tissues, leading to ischemia, chronic organ damage, and eventual tissue death and organ failure. Several narrative reviews have detailed the array of complications associated with SCD, including but not limited to neurological, cardiovascular, developmental, obstetric, ocular, bone, dermatological, renal, and infectious complications.[16–19] Additionally, socioeconomic factors such as access to and utilization of affordable health care, and environmental factors like climate and air quality have been associated with the frequency and severity of SCD complications.[2,20–22]

The lack of a common understanding of SCD complications negatively affects patients’ medical care, well-being, and survival. In particular, it is important to understand how the risk of complications varies over the lifespan, as patients may be especially vulnerable to lapses in treatment or preventative care in the transition from pediatric to adult care.[23–25] Using health-care utilization data among commercially insured individuals in the USA from 2016 to 2019, this study aimed to estimate the prevalence and incidence (per person-year) as well as the relative risks or incidence rate ratios for chronic and acute disease complications among individuals with SCD compared to those without SCD, overall and stratified by age and sex. We frame these estimates deliberately as exposure-outcome effect sizes rather than as a US descriptive profile because such effect sizes are the quantity that burden-of-disease frameworks require in order to attribute non-hematologic disease to SCD. The meta-regression design allows for future data additions from other sources that can measure comparative risk from parts of the world with the highest prevalence of SCD.

## Methods

### Data sources

This study was completed as part of Global Burden of Diseases, Injuries, and Risk Factors Study which was reviewed and approved for waiver as non-human subjects research by the University of Washington Institutional Review Board (study number 9060). The requirement for informed consent was waived because the data were fully de-identified. Longitudinal health-care utilization and demographic information for years 2016 through 2019 were obtained from Truven MarketScan, a national health insurance claims database with de-identified state-level data for over 50 million beneficiaries with employer-sponsored health insurance.[26] Specifically, this included: (1) commercial claims and encounters database for privately insured and employer-insured persons and their dependents in the USA (by state) and (2) the Medicare supplemental and coordination of benefits database for Medicare-eligible retirees with employer-sponsored supplemental plans (also separate by state). This represents a convenience sample of the population of employed or formerly-employed individuals and their dependents in the USA. Non-physician encounters provided in the laboratory or pharmacy setting were excluded from outpatient services. For each facility (outpatient versus inpatient) and reporting year, we deduplicated data rows with identical patient identifier, service date, diagnosis position, and ICD-10 code.

### Inclusion and exclusion criteria

All records with at least six months of continuous insurance enrollment across calendar years 2016 through 2019 were included. A minimum six-month follow-up was chosen to ensure sufficient data availability for each observation. There were no exclusion criteria related to pharmaceutical interventions. Person-time accrued while a beneficiary resided in a US territory was excluded, but individuals could still contribute person-time from calendar years in which they resided in a US state.

### Identifying SCD cases and controls

Our primary exposure was SCD diagnosis. An SCD diagnosis was defined as having at least one inpatient visit or two outpatient visits with any derivatives of ICD-10 code D57, excluding D57.3 (sickle cell trait), an approach that aligns with previous studies of SCD using insurance claims data.[27,28] An SCD diagnosis at any point during the study window qualified a patient as an SCD cohort member. To maximize the utilization of Marketscan, we also assigned each individual to a specific genotype – HbSS, HbSC, HbSβ+/HbSβ, and HbOS (all others) – with an algorithm adjudicating diagnosis disagreements. [6] Of note, however, because genotype assignment based on ICD-10 codes has not been validated these results are only shown in Supporting Information.

Patients in the control group (i.e., without SCD) were excluded in all analyses if they had chronic conditions similar to SCD by the mechanism of anemia (hemolysis). ICD-10 codes for exposure and control definitions are defined in S1 Appendix.

In addition to dichotomous assignment to the SCD or control group, we grouped all records by age strata (<20, 20–29, 30–39, 40–49, 50–59, and 60+ years), sex (male, female), and state of residence at enrollment into insurance program. The MarketScan database did not contain information on socioeconomic status, race, or ethnicity.

### Outcome definitions

A total of 30 candidate outcomes were evaluated for association with SCD, including known direct, disease-defining sequelae of SCD (e.g., vaso-occlusive crises, stroke) and other conditions where SCD is hypothesized or has been reported to be a risk factor (e.g., kidney disorders, respiratory diseases, pregnancy complications). The MarketScan database did not contain mortality information and there were not enough data on certain complications of SCD (e.g., leg ulcer, acute chest syndrome) to perform analyses. Because anemia is a key feature of SCD and can improve after a transfusion, which we were unable to measure with our database, we did not include this outcome in our analysis.

Outcome-specific ICD-10 codes were informed by case-finding algorithms in published literature and are defined in S1 Appendix. For chronic disease outcomes, a diagnosis was defined as the presence of relevant ICD-10 codes in at least one inpatient visit or two outpatient visits on different service dates at any point during the follow-up. Following the initial diagnosis, the chronic disease was assumed present through the end of follow-up. For acute disease outcomes, a diagnosis was defined as the presence of relevant ICD-10 codes in at least one inpatient or one outpatient visit within a defined duration window (i.e., the duration of time following the first incidence of the acute diagnosis after which another diagnosis may reflect a new incident case for the same individual rather than a subsequent visit for the same disease episode). Multiple diagnosed cases could occur across the follow-up period. Diseases of pregnancy were measured among all women of reproductive age (i.e., aged 10-54 years), though estimates for females under age 20 and age 50 or older were censored due to small counts. Use of disease-modifying treatments, such as hydroxyurea, transfusions, or dialysis, was not measured as part of this study.

### Data preparation and statistical modelling

Rate (prevalence or incidence) and ratio (relative risk or incidence rate ratio) models were fit independently. We calculated prevalence and incidence, respectively, as the number of diagnosed cases over the total number of individuals or person-years, where the presence of at least one diagnosis in a year was counted as one person-year, in the exposed or unexposed groups. A number of processing steps were completed prior to meta-regression. First, to avoid bias introduced by small sample sizes and zero counts in log- and logit-transformed data, we applied an offset to incidence and prevalence data, replacing zero prevalence or incidence rates with 0.5 times the lowest non-zero value, and replacing prevalence of one with one minus 0.5 times the highest non-zero value. We calculated the standard error of prevalence using the Wilson method and of incidence using the Poisson distribution method, considering the number of reported cases.[29–32] The input data for RR and IRR models were calculated from the prevalence or incidence observations with SCD cases as numerator and controls as denominator. Certain outcomes were excluded from reporting if the number of all-age, both-sex cases for that group totaled fewer than 50 or, for ratios, if the all-age, both-sex rate in the exposed or unexposed group was less than 5 per 10,000.

The observations of age-, sex-, and state-specific prevalence and RRs (for chronic outcomes) and incidence rates and IRRs (for acute outcomes) were inputs to outcome-specific meta-regression with Bayesian priors, regularization, and trimming (MR-BRT) models, whose methodology has been detailed elsewhere.[33] For each SCD complication, we conducted three MR-BRT models with random slopes and intercepts for different states: (1) without any additional covariates (i.e., a “conventional” meta-analysis), (2) with sex as a categorical covariate, and (3) with sex and age as categorical covariates (i.e., allowing for non-linear age patterns). In the meta-regression models, each effect size was weighted by the inverse of its variance, leading to estimates with smaller uncertainty having more weight in the models. Each MR-BRT model yielded 500 random samples (assuming a normal distribution) using the point estimate and the uncertainty from both fixed and random effects.

### Model evaluation and uncertainty

One of the characteristic strengths of MR-BRT is its ability to quantify uncertainty both using conventional uncertainty intervals as well as by incorporating the additional quantitative uncertainty arising from residual uncertainty of the input data and model fit. Incorporation of residual heterogeneity into assessing model fits serves as a powerful mechanism to evaluate overall strength of evidence supporting associations between exposures and outcomes. All uncertainty intervals shown in our results include residual heterogeneity.

The delta method was used to transform standard errors of a logit-transformed prevalence or log-transformed rate back to the linear scale.[34] The 2.5th, 50th, and 97.5th percentiles were calculated to represent the median and 95% uncertainty intervals (UI). All statistical analyses were performed using R version 4.2.2.[35] We used Claude (Anthropic) to assist with two aspects of manuscript preparation: (1) generating and formatting the code syntax used to produce Tables 1–3, and (2) copy-editing for grammar, style, and readability. The tool was not used for study design, data collection, analysis, interpretation of results, or generation of original text. The authors reviewed, verified, and edited all AI-assisted output and take full responsibility for the content of the manuscript. This study is reported in accordance with the STROBE statement for observational research; the completed checklist is provided as Supplemental Table 3. We additionally report against the Guidelines for Accurate and Transparent Health Estimates Reporting (GATHER; Supplemental Table 1).Results

**Table 1.** Characteristics of the eligible study population (N=52,709,530)

|  | **SCD N (%)** | **Non-SCD\* N (%)** |
|---|---|---|
| Eligible individuals | 15,462 | 52,694,068 |
| Females | 9,259 (60) | 27,357,304 (52) |
| *Age distribution* |  |  |
| <20 years | 4,351 (28) | 12,201,085 (23) |
| 20 to 29 | 2,938 (19) | 8,648,232 (16) |
| 30 to 39 | 2,644 (17) | 8,002,530 (15) |
| 40 to 49 | 2,308 (15) | 8,061,858 (15) |
| 50 to 59 | 1,859 (12) | 8,590,249 (16) |
| 60 plus | 1,362 (9) | 7,190,114 (14) |
| *Genotype distribution* |  |  |
| HbSS | 11,496 (74) | ·· |
| HbSC | 2,293 (15) | ·· |
| HbSβ+/0Thal | 1,329 (9) | ·· |
| HbS-Other | 344 (2) | ·· |

*\*Eligible individuals in the Non-SCD group do not have SCD, SCT nor any other hemoglobinopathy or hemolytic anaemia per MarketScan data*

**Table 2.**
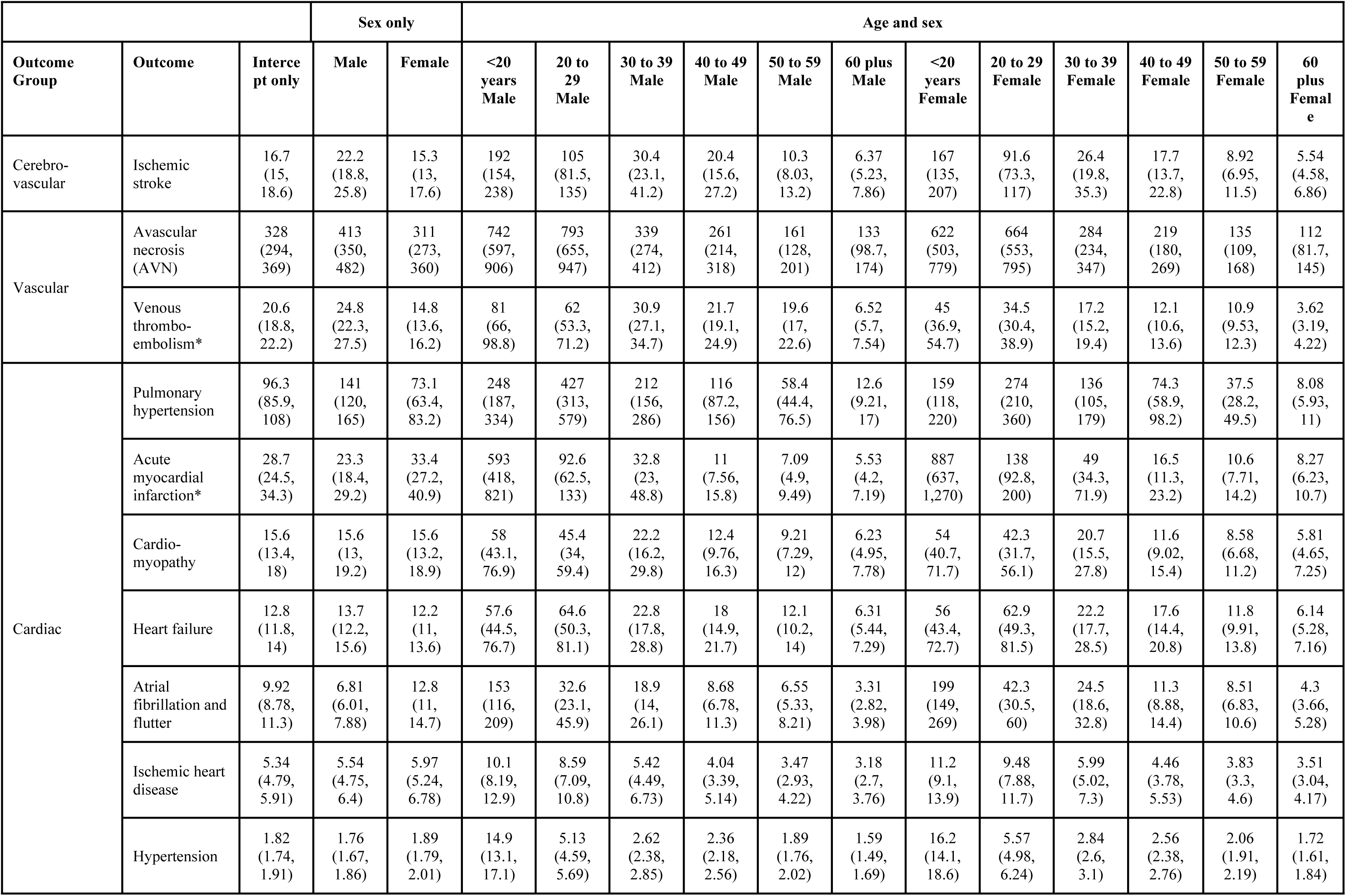
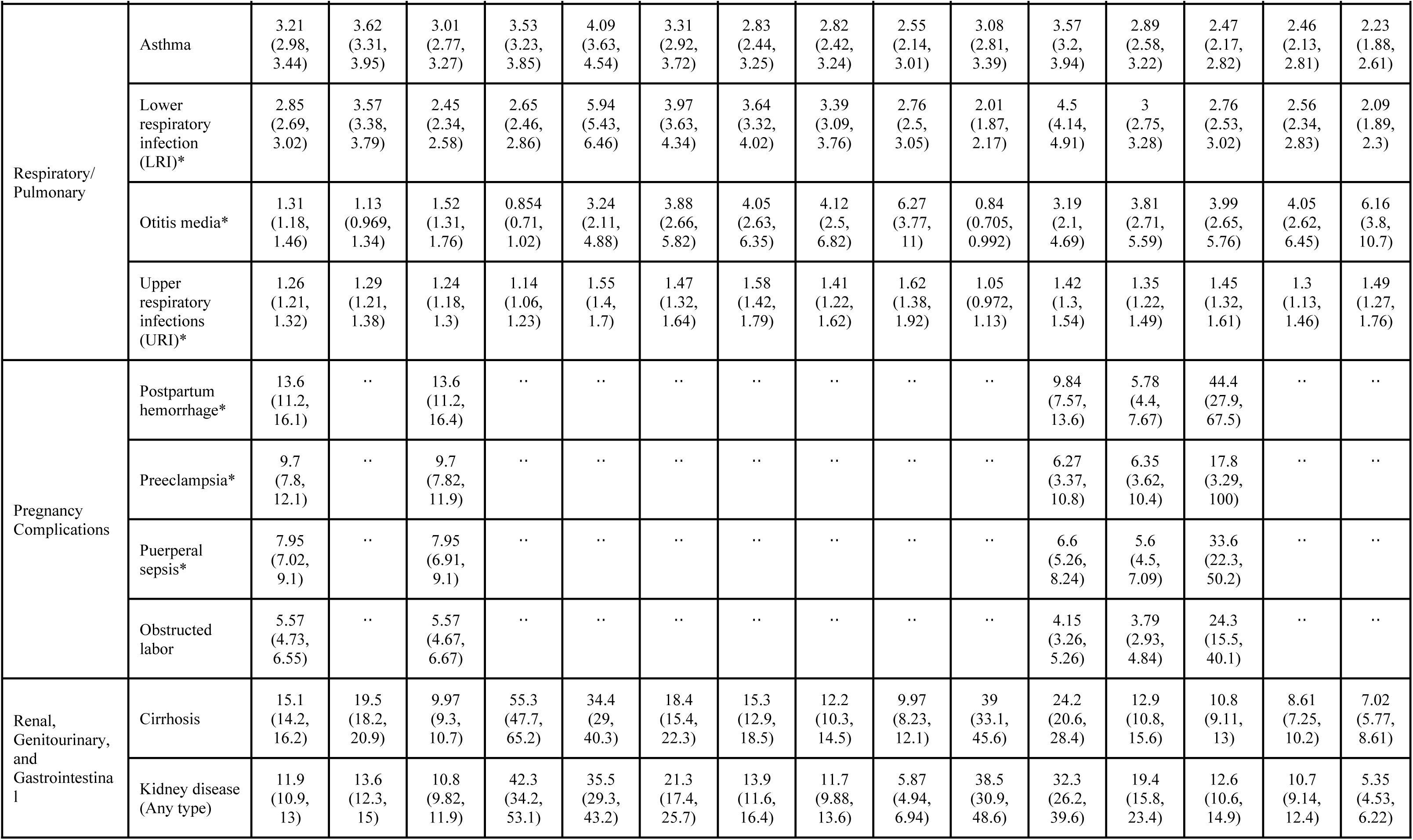
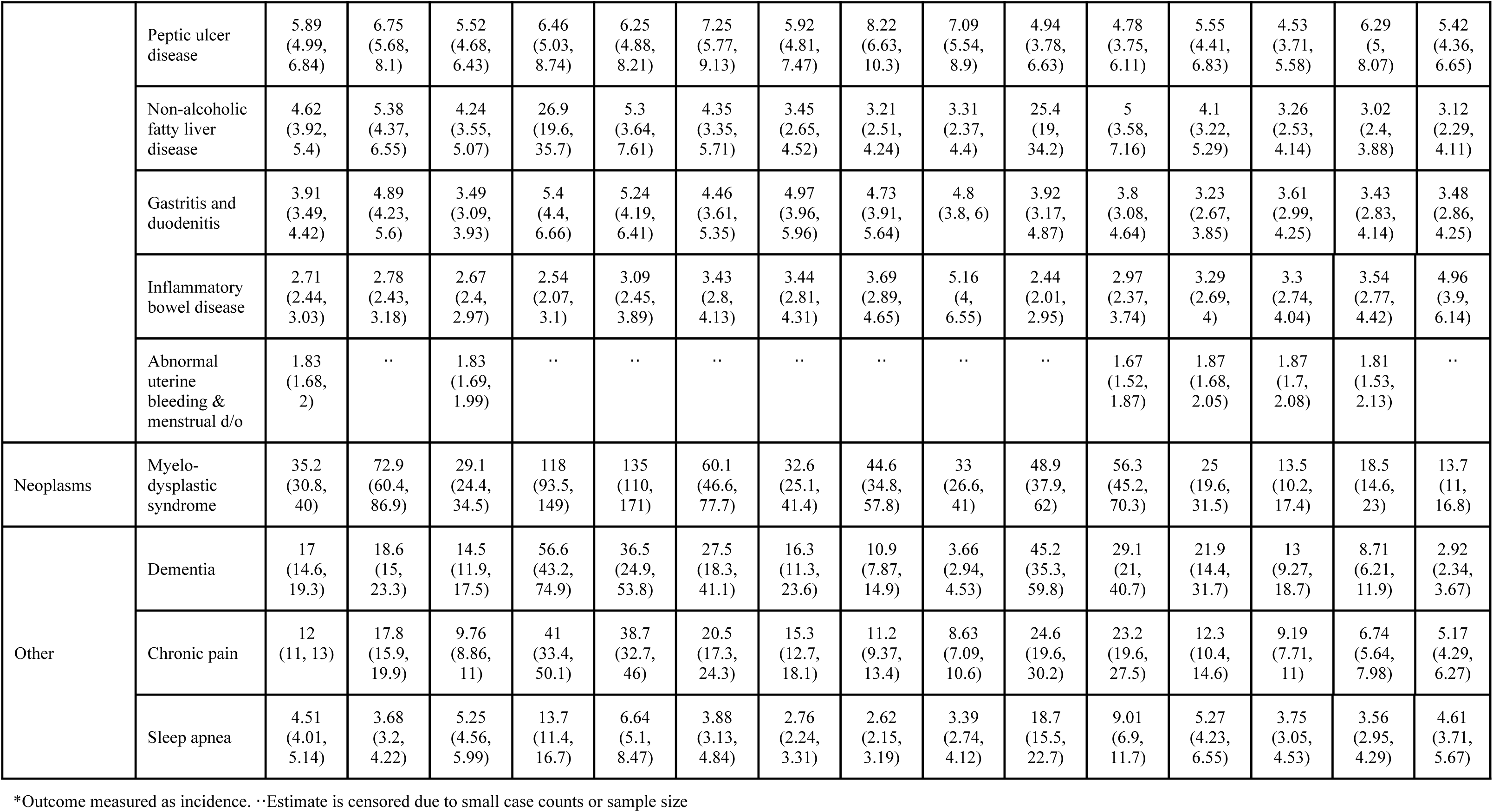
Relative risk and incidence rate ratio of health outcomes in patients with versus without sickle cell disease, by age and sex, 2016-2019 USA.

**Table 3.**
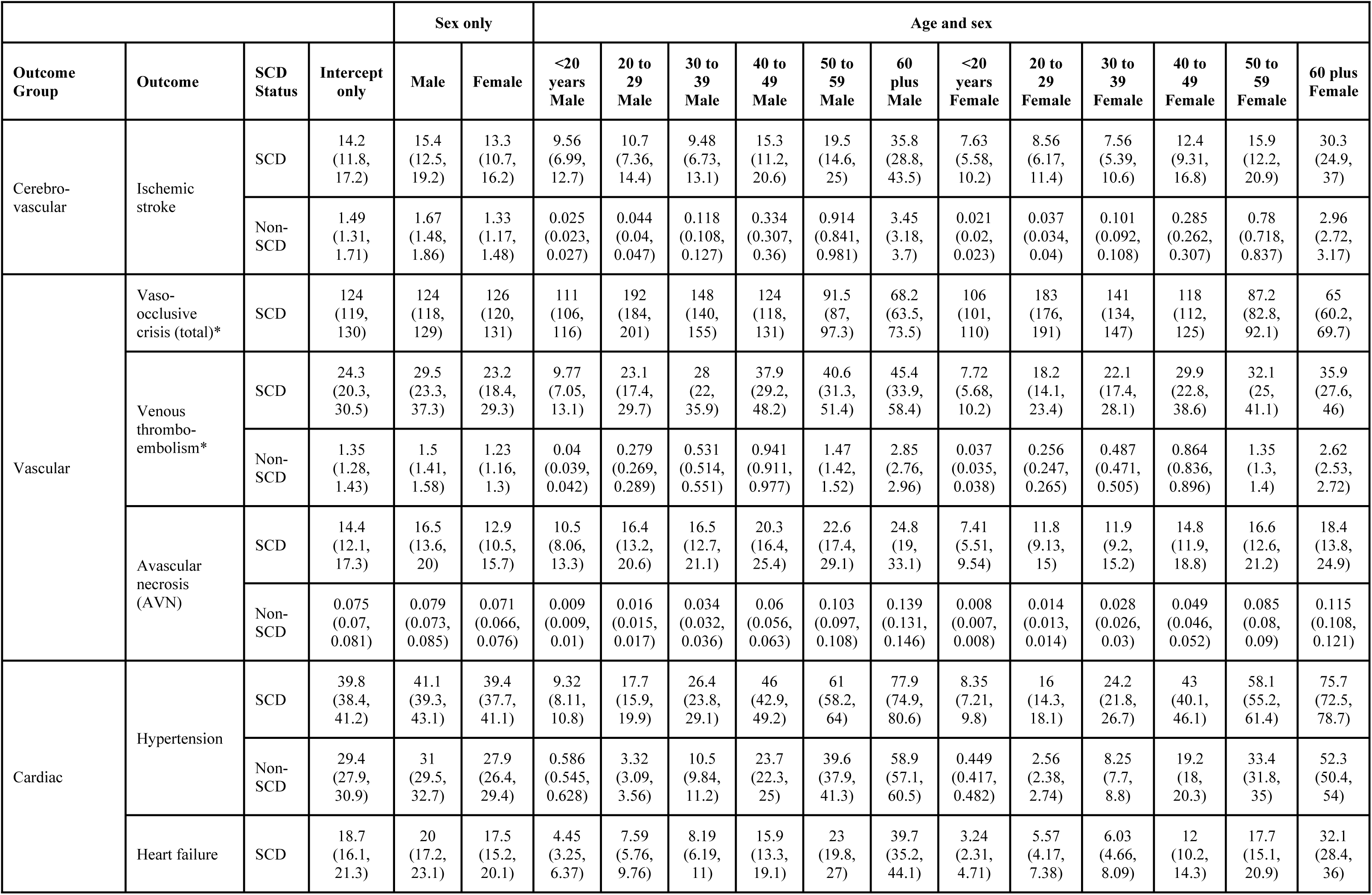
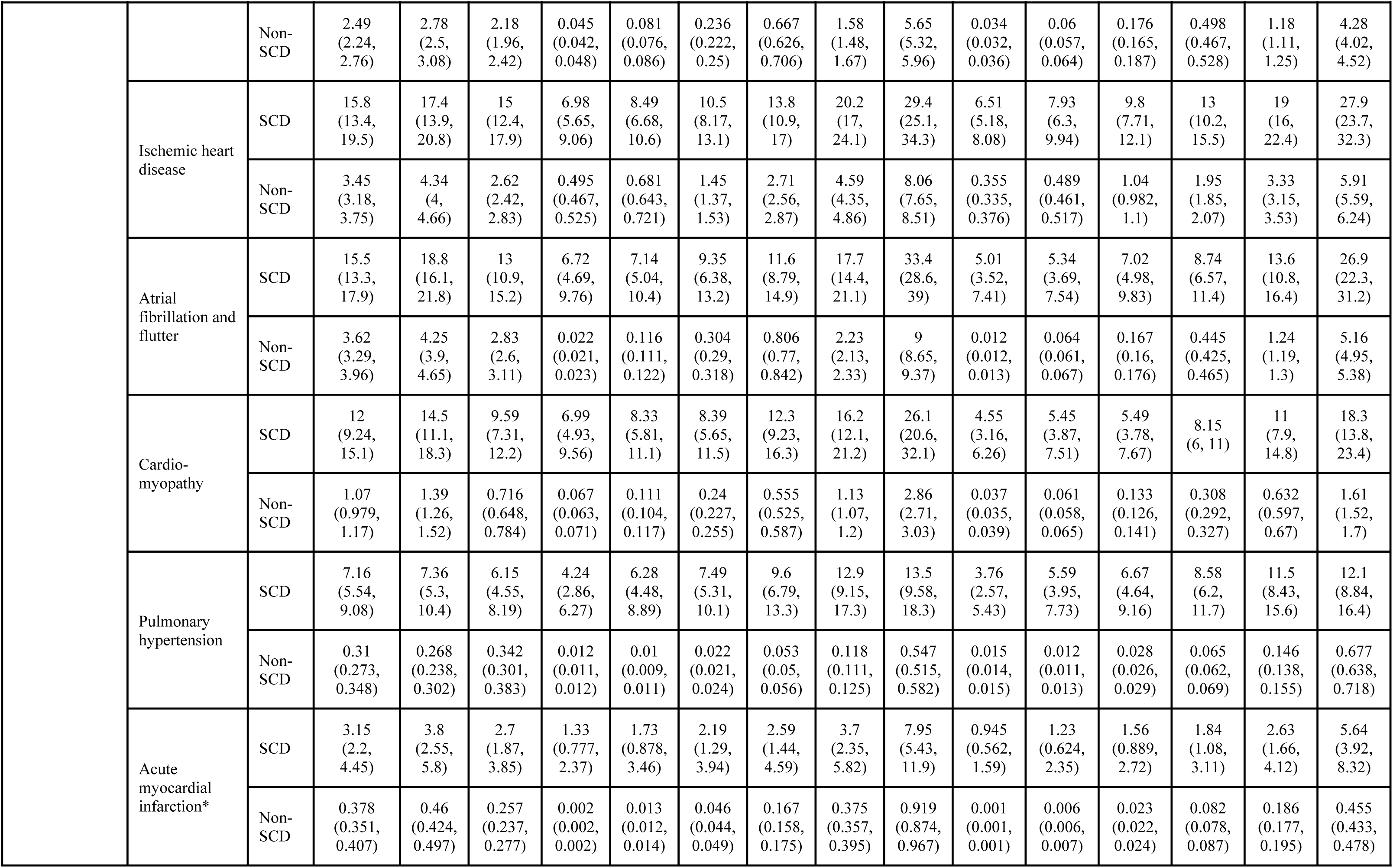
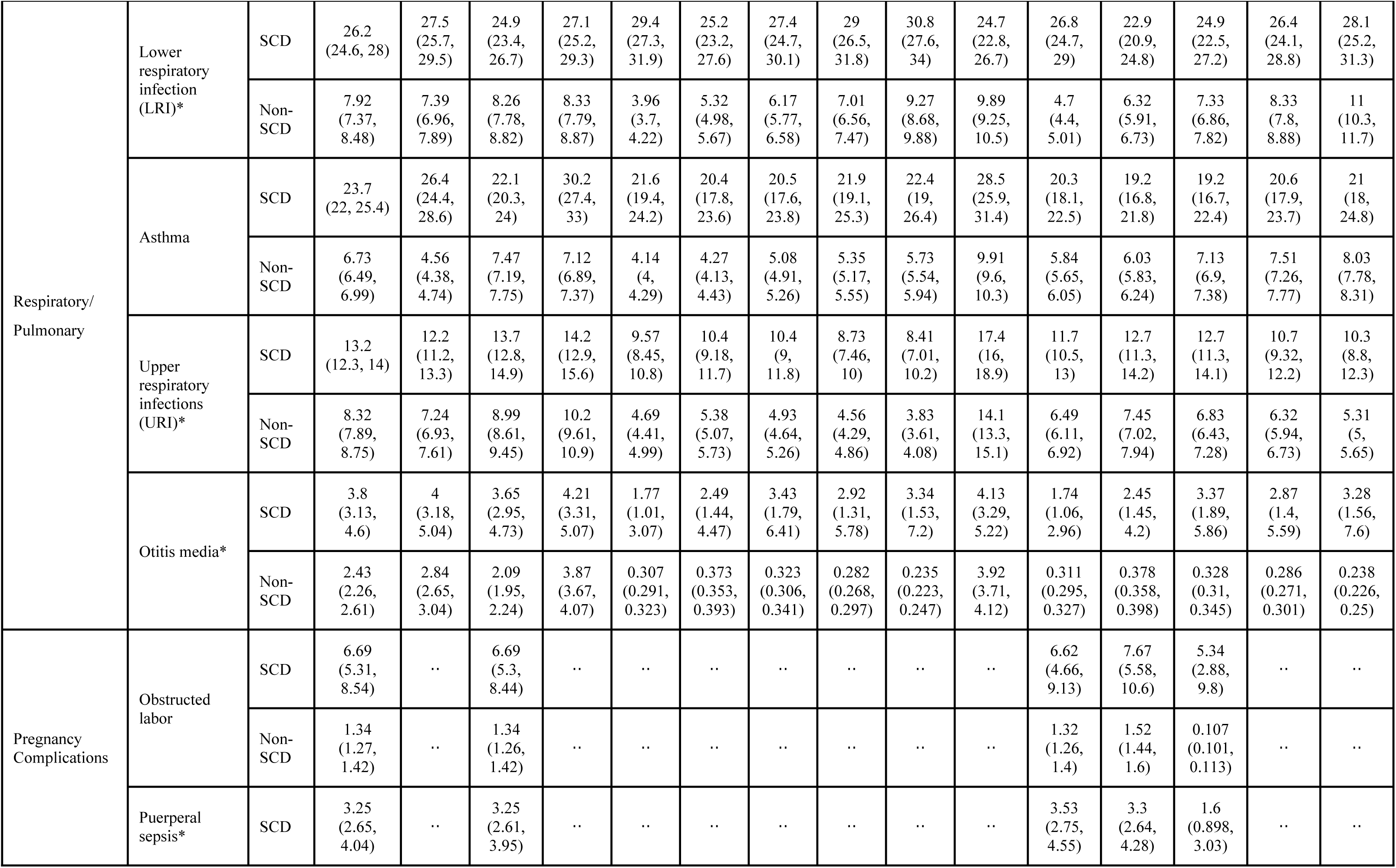
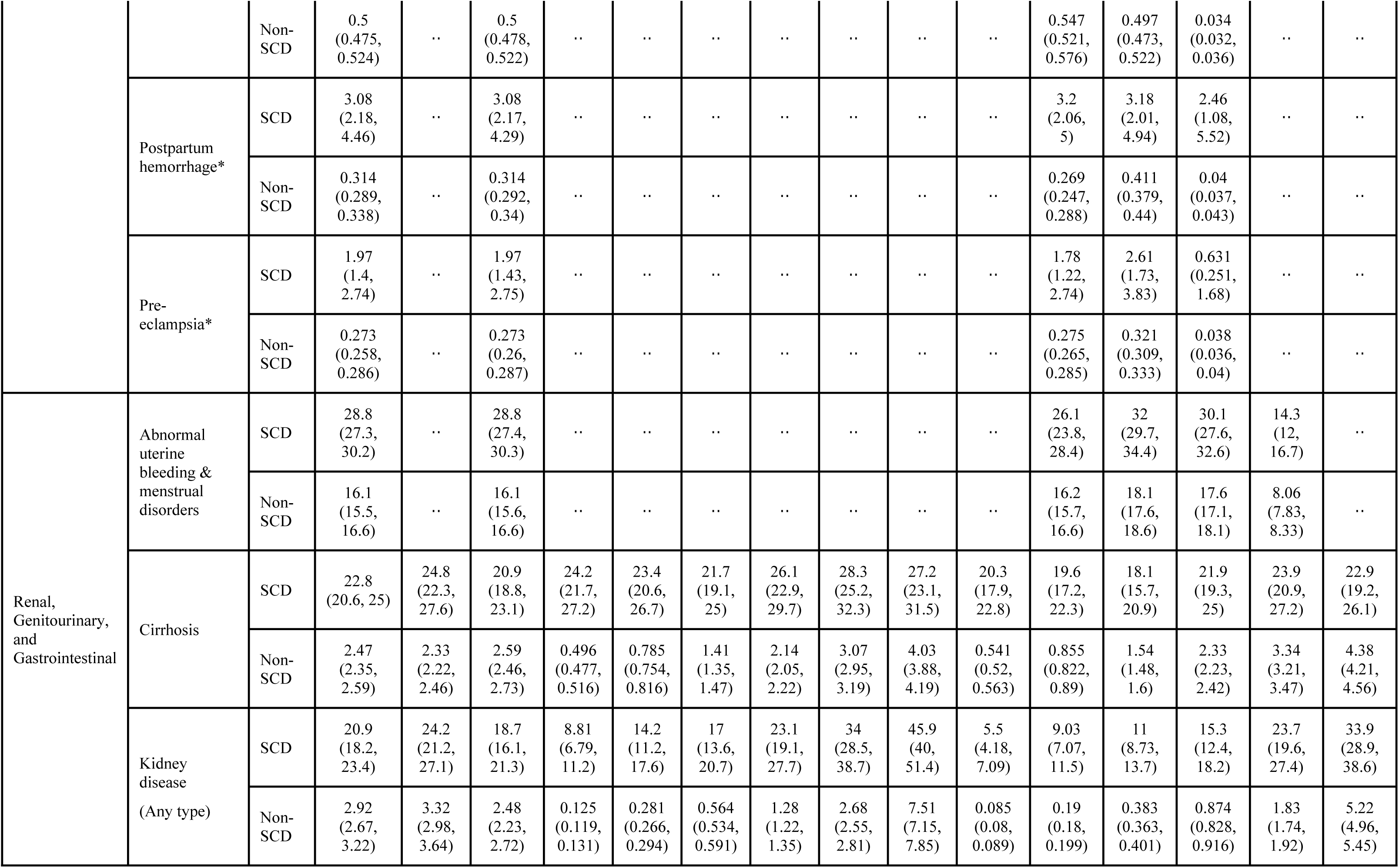
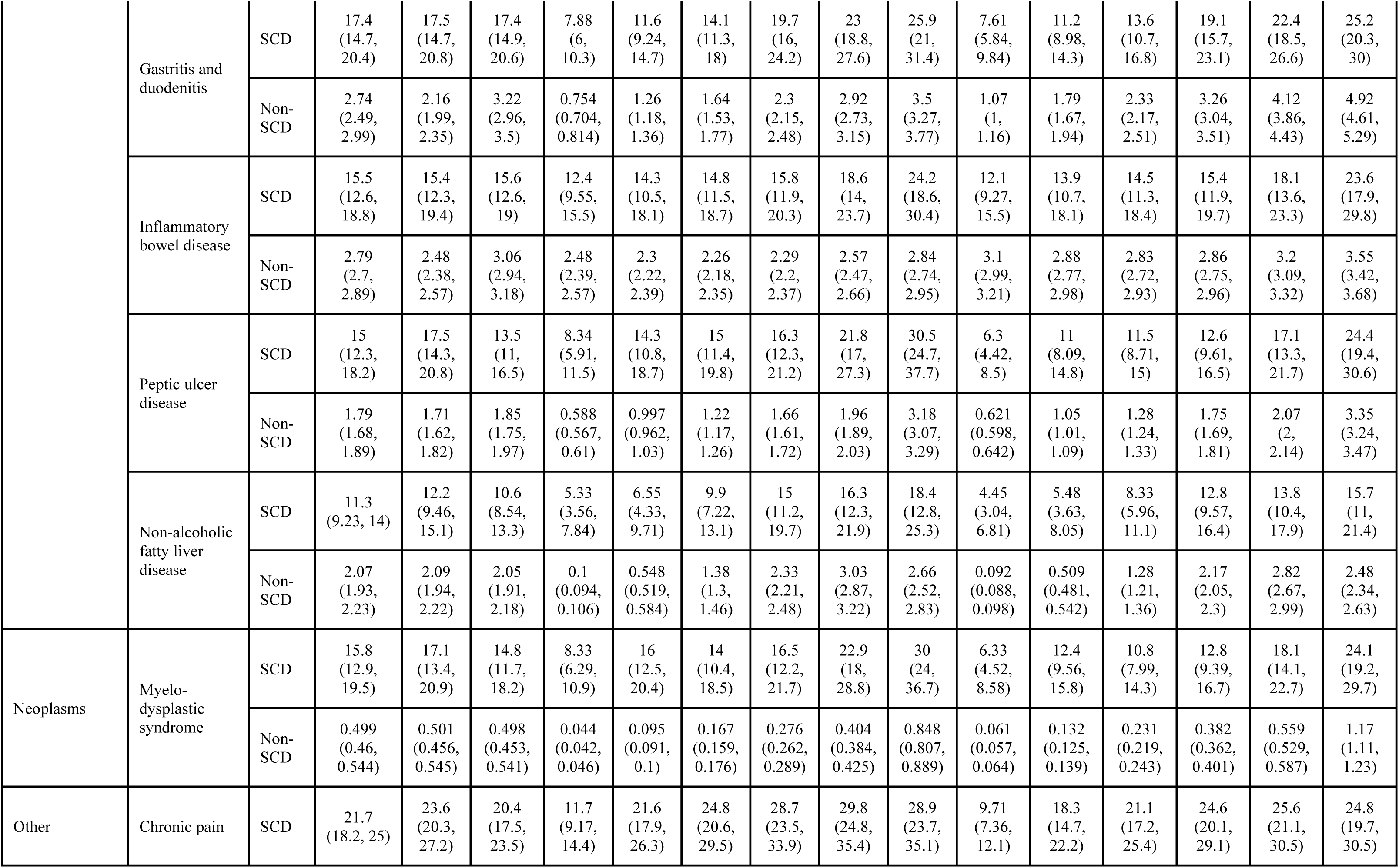
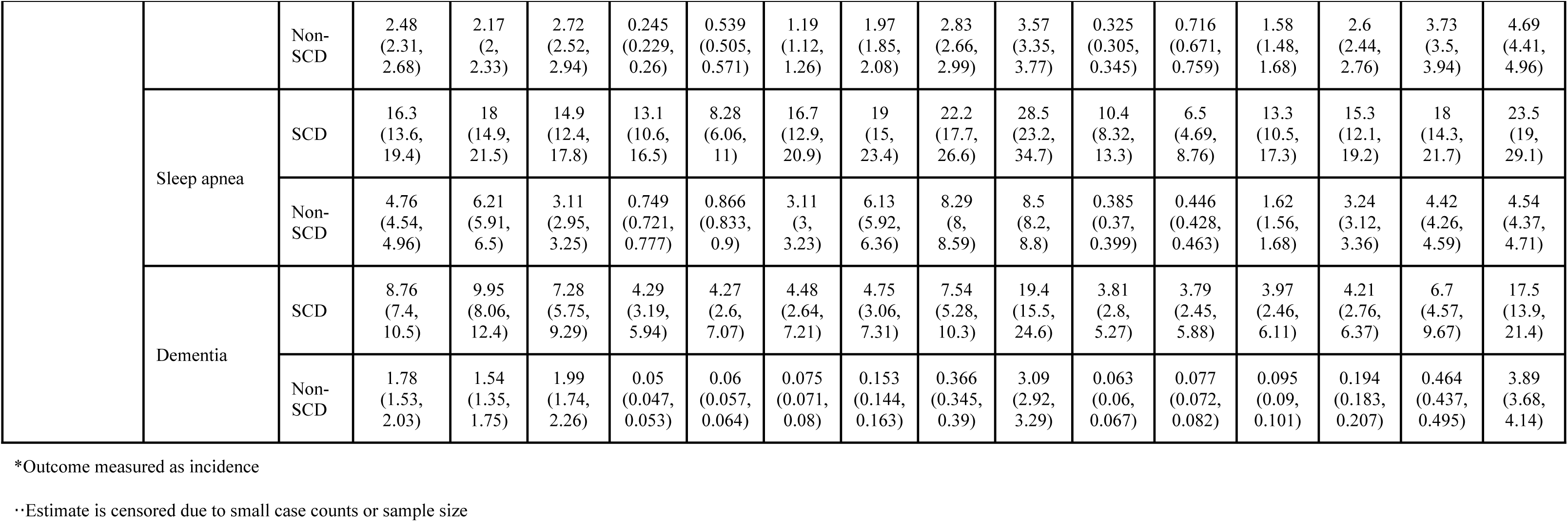
Prevalence [per 100 persons] and incidence [per 100 person-years] (with 95% uncertainty interval) of health outcomes by SCD status, age, and sex, 2016-2019 USA.

## Results

Of all insurance beneficiaries in the MarketScan database, 52,709,530 had at least six months of continuous enrollment, resided in the USA, and did not have other hemoglobinopathies. There were a total of 15,462 individuals with SCD and 52,694,068 individuals without SCD or other hemoglobinopathies. Both groups had more females than males, with 60% and 52% of the SCD and control cohorts being female. Our genotype assignment algorithm identified (74%) of the SCD cohort as having the HbSS genotype (Table 1, Note: all additional genotype-specific results are shown only in Supporting Information).

In intercept-only meta-analyses, SCD had a significantly elevated RR or IRR for all included health outcomes (Fig 1, Table 2). The most elevated RRs and IRRs were in chronic and acute outcomes, respectively, that are very rare in the general population but are known complications of SCD, such as pulmonary hypertension (RR=96.3, 95% UI 85.9–108.1) and avascular necrosis (IRR=328, 294-369)

**Fig 1.**
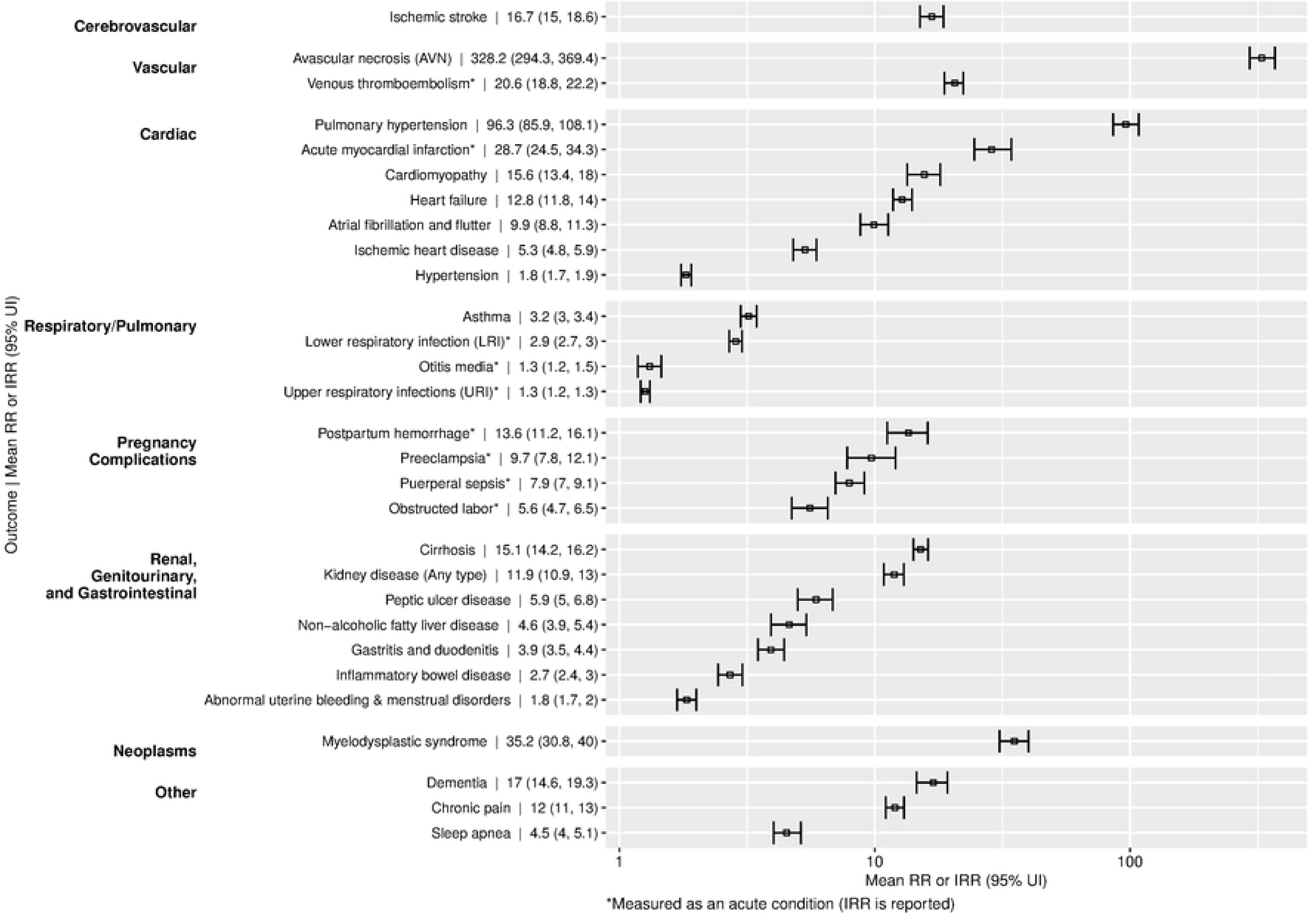
Relative risk and incidence rate ratio of health outcomes in patients with and without sickle cell disease, 2016−2019 USA This figure shows pooled all age and sex relative risk (RR; for chronic conditions) and incidence rate ratio (IRR; for short-term/ acute conditions) in SCD versus non-SCD individuals. The mean and 95% uncertainty intervals are plotted in log (base 10) scale. Outcomes marked with * were measured as acute conditions and therefore reported as IRR. Abbreviations: RR = relative risk; IRR = incidence rate ratio.

This analysis also found that there were significantly elevated risks for other acute and chronic cardiovascular conditions, such as acute myocardial infarction (IRR=28.7, 24.5–34.3), and venous thromboembolisms (IRR=20.6, 18.8–22.2). Additionally, individuals with SCD had significantly elevated risks of kidney disease (RR=11.9, 10.9–13.0), preeclampsia (IRR=9.7, 7.8–12.1), and cirrhosis (RR=15.1, 14.2–16.2). The lowest estimated IRR via the intercept-only models was for upper respiratory infections but still had a 26% higher risk in individuals with SCD versus without SCD (IRR=1.26, 95% UI 1.21–1.32).

Sex- and age-specific results revealed important pattens. (Table 2, Fig 2, Fig 3, eFigure 1-3). In the SCD cohort, vaso-occlusive crises and hypertension had the highest incidence and prevalence, respectively, amongst males (124 events per 100 person-years [95% UI 118–139] and 41.1% [39.3%–43.1%]) and females (126 events per 100 person-years [120–131] and 39.4% [37.7%–41.1%]) (Table 3). The age pattern of prevalence and incidence of most conditions revealed monotonic increases in both SCD and non-SCD groups, but the age trend was much more stark amongst those without SCD or other hemoglobinopathies.

**Fig 2.**
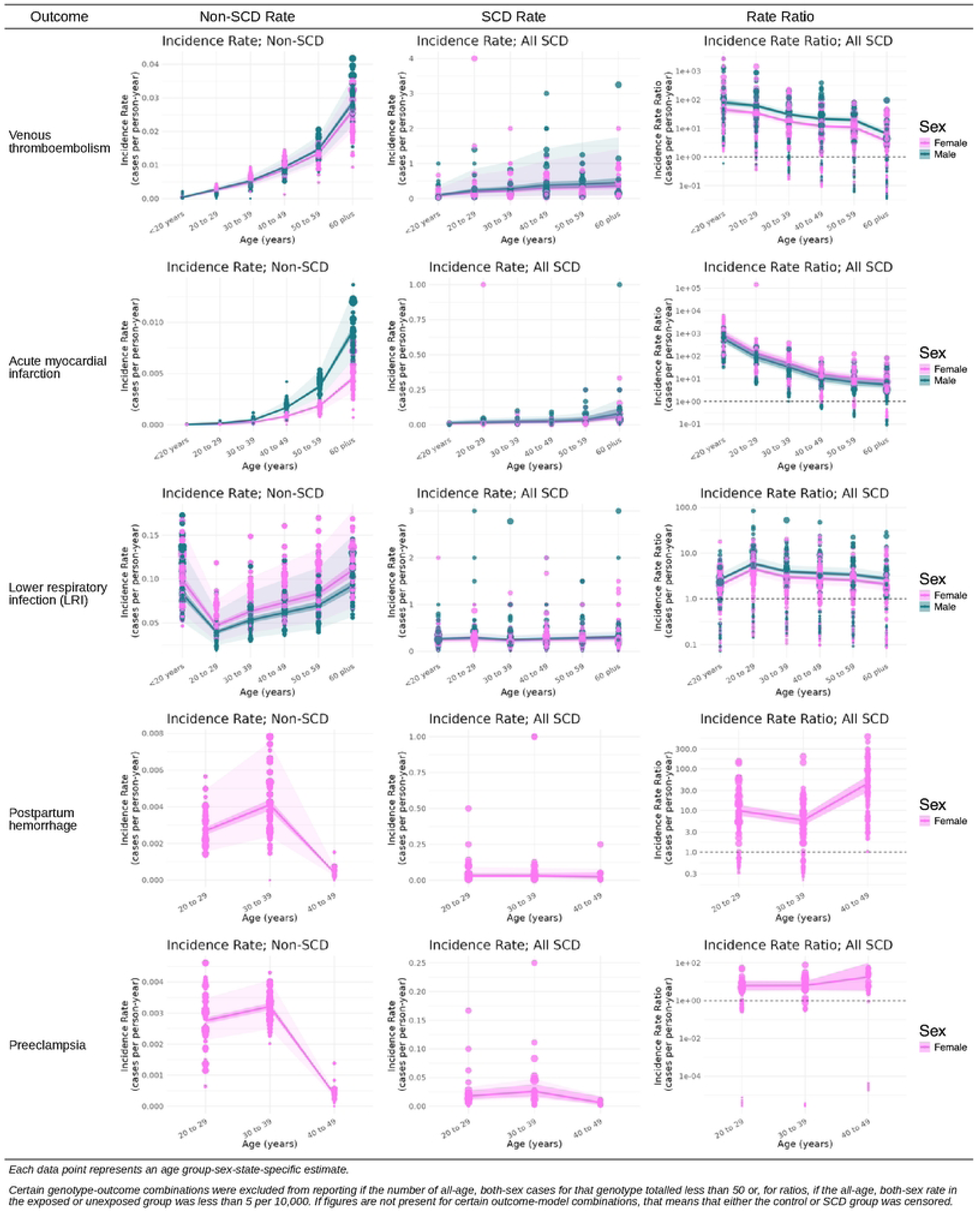
Comparative prevalence and incidence of acute health complications among people with and without sickle cell disease, by age and sex, 2016-2019, USA This figure highlights age- and sex-specific incidence rate ratio (IRR; for short-term/ acute conditions) in SCD versus non-SCD individuals for selected outcomes including venous thromboembolism, acute myocardial infarction, lower respiratory infections, postpartum hemorrhage, and preeclampsia. The mean and 95% uncertainty intervals are plotted for males (blue) and females (pink). Each data point represents an age group-sex-state-specific estimate.Certain genotype-outcome combinations were excluded from reporting if the number of all-age, both-sex cases for that genotype totalled less than 50 or, for ratios, if the all-age, both-sex rate in the exposed or unexposed group was less than 5 per 10,000. If figures are not present for certain outcome-model combinations, that means that either the control or SCD group was censored. Abbreviations: RR = relative risk; IRR = incidence rate ratio; SCD = sickle cell disease.

**Fig 3.**
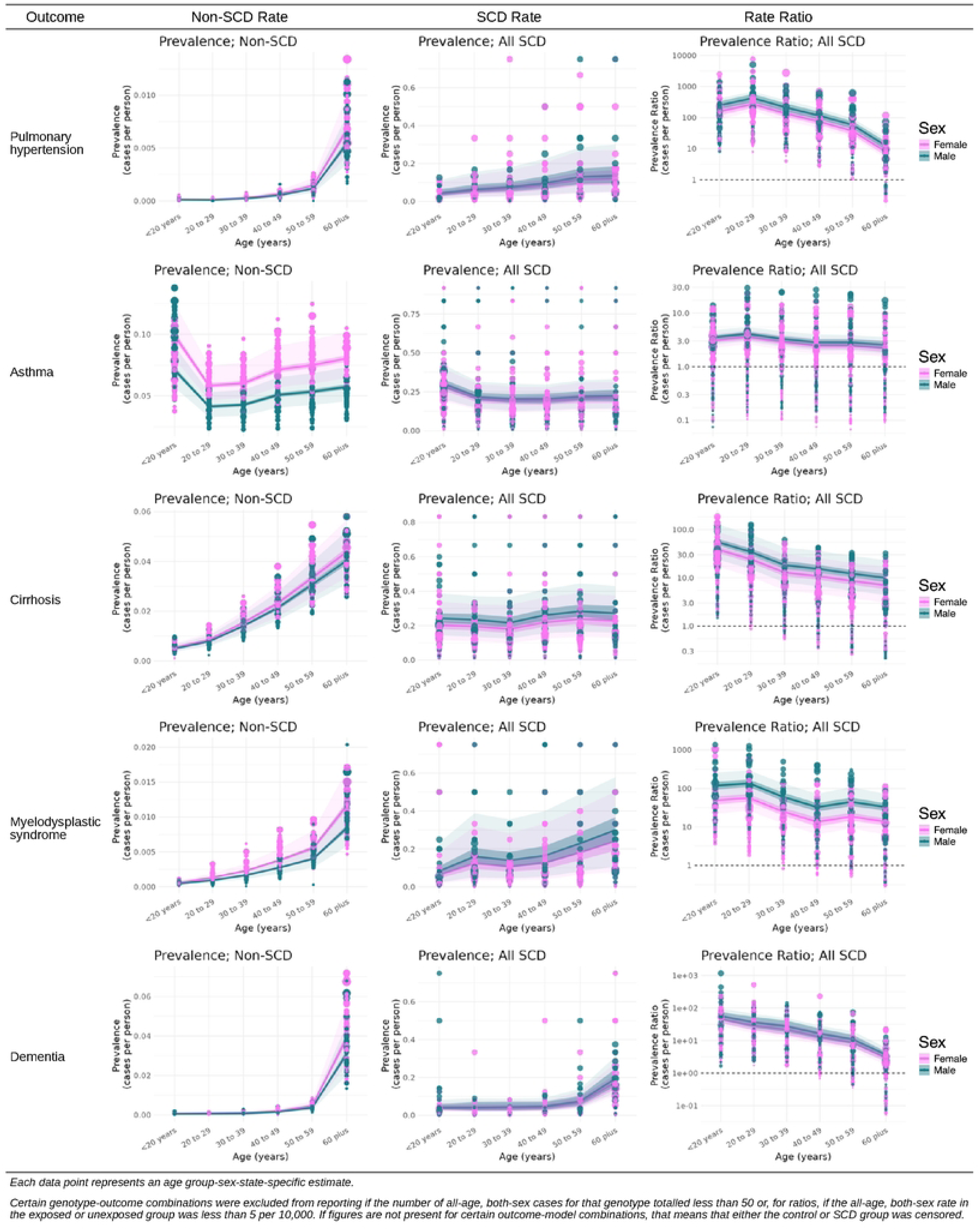
Comparative prevalence and incidence of chronic health complications among people with and without sickle cell disease, by age and sex, 2016-2019, USA This figure highlights age- and sex-specific relative risk (RR; for long-term/ chronic conditions) in SCD versus non-SCD individuals for selected outcomes including pulmonary hypertension, asthma, cirrhosis, myelodysplastic syndrome, and dementia. The mean and 95% uncertainty intervals are plotted for males (blue) and females (pink). Each data point represents an age group-sex-state-specific estimate.Certain genotype-outcome combinations were excluded from reporting if the number of all-age, both-sex cases for that genotype totalled less than 50 or, for ratios, if the all-age, both-sex rate in the exposed or unexposed group was less than 5 per 10,000. If figures are not present for certain outcome-model combinations, that means that either the control or SCD group was censored. Abbreviations: RR = relative risk; IRR = incidence rate ratio; SCD = sickle cell disease.

The comparatively “flat” age trends in patients with SCD, reflecting early onset of many conditions, meant that the highest mean RRs or IRRs among both males and females for 14 of 24 measured conditions were in the under-20 age group. Under-20 females and males with SCD, respectively, had 24.6 (19.6–30.2) and 41.0 (33.4–50.1) times the risk of chronic pain and 167 (135–207) and 192 (154–238) times the risk of ischemic stroke compared to patients without SCD in the same age group (Table 2). The IRRs and RRs of otitis media, inflammatory bowel disease (IBD), and upper respiratory infections (URIs) were highest in individuals 60 years of age and older among both males and females. Male patients ages 60 years and older with SCD had 6.27 (95% UI 3.77–11.0) times the incidence rate of otitis media, 5.16 (4.00–6.55) times the risk of IBD, and 1.62 (1.38–1.92) times the rate of URIs compared to male patients aged 60 years and older without SCD or other hemoglobinopathies (Table 2). Female patients aged 60 years and older with SCD had 6.16 (3.80–10.7) times the incidence rate of otitis media, 4.96 (3.90–6.10) times the risk of IBD, and 1.49 (1.27–1.76) times the rate of upper respiratory infections compared to female patients aged 60 years and older without SCD or other hemoglobinopathies (Table 2).

Rates and ratios differed significantly by sex for half of the outcomes measured. Pregnant females with SCD had incidence rates 5.6 to 13.6 times higher for health complications compared to pregnant females without SCD or other hemoglobinopathies (Fig 1). For most maternal outcomes, incidence rates were highest in individuals with or without SCD aged 30 to 39 years (Fig 3, Table 3). The rates of asthma and atrial fibrillation were higher in males with SCD compared to females with SCD (26.4 [24.4–28.6] versus 22.1 [20.3–24.0] and 18.8 [16.1–21.8] versus 13.0 [10.9–15.2]). The RRs were statistically significantly more elevated in females compared to males for only two conditions: atrial fibrillation (12.8 [11.0–14.7] for females versus 6.81 [6.01–7.88] for males) and sleep apnea (5.25 [4.56–5.99] for females versus 3.68 [3.20–4.22] for males).

## Discussion

We identified significantly elevated rates of numerous adverse health outcomes in a commercially insured cohort of patients with SCD living in the USA, including not only established SCD complications, but also other common illnesses where SCD is a biologically plausible risk factor. SCD can affect nearly every organ system, and males and females with SCD are at a significantly elevated risk of health complications, including potentially fatal cardiac and vascular events, at all ages. Though risk of cardiac and vascular events increases with age in patients with and without SCD, the mean relative risks and rate ratios of complications in those younger than 20 years were extremely elevated in part due to the rarity of such diseases in children and young adults without SCD or other hemoglobinopathies and may indicate the need for increased pediatric surveillance strategies. The evidence presented in this study, alongside factors associated with increased mortality such as socioeconomic status, household composition and disability, minority status and primary language, housing type, and access to transportation, should be considered in patient case management and education.[36]

Our findings are consistent with evidence in the literature showing that the overall disease frequency among patients with SCD is notably higher than in those without SCD.[22,37,38] Studies indicate that the increased occurrence of complications in SCD is due to the multi-system impact of the disease, driven by vaso-occlusive crises (VOCs), chronic hemolysis, and resulting end-organ damage, emphasizing the need for an expert, interdisciplinary care team from an early age.[39–41] Few studies report the RR or IRR between patients with versus without SCD. In a retrospective cohort study using a multisystem database with over 100 million patients across 30 countries, patients with SCD had higher risk of many cardiovascular outcomes including ischemic heart disease, atrial fibrillation and flutter, heart failure, and pulmonary hypertension, even after adjusting for demographic and cardiovascular risk factors.[42] In a meta-analysis, Oteng-Ntim and colleagues found that pregnant women with SCD have high risks of adverse maternal and perinatal outcomes, which is consistent with our findings.[43] A meta-analysis of three studies by Noubiap and colleagues also reports increased odds of venous thromboembolism in non-pregnant adults with SCD (OR = 4.43 [2.62–7.48]), though at a lower magnitude than the all-age, both-sex IRR estimated in our study (IRR=20.6 [95% UI 18.8–22.2]), likely due to the different study populations and sample sizes.[44] Consistent with our findings, a study by Manwani and colleagues investigating the complications associated with SCD using US administrative claims data from the Optum Research Database, ICD-coded diagnoses, and employing similar inclusion criteria for identifying cases, found that SCD patients experience a high burden of complications, including acute kidney injury, chronic kidney disease, and stroke, which increase with age.[45] There are limited and mixed reports on myelodysplastic syndrome (MDS) in patients with SCD, but the characteristic chronic hemolysis and subsequent inflammation in SCD provides a potential mechanism of oncogenesis worth monitoring in this population.[46] Another potential explanation for elevated rates of MDS in patients with SCD could be miscoding ICD-coded diagnoses as patients with SCD generally have more hematology encounters than an average control patient.

This study is not without limitations. First, while the use of ICD-coded data is a gold standard of population health measurement and usage of administrative data allows for efficient matching to control patients, ICD-coded data are not perfect.[47] In the USA, provision and documentation of medical care is typically siloed from coding and billing which, coupled with complexities of fragmented documentation platforms, incomplete provider knowledge about SCD, and presence of multiple comorbidities, can lead to omission of codes and/or misassignment of SCD status, especially for less severe complications or when care is provided by non-specialists.[27,48] We aimed to mitigate by prioritizing care setting and provider specialty reports for SCD status and by requiring at least one inpatient and two outpatient encounters for chronic disease and duration windows for acute conditions. Second, while MarketScan data has advantages in terms of sample size, it’s reliance on commercial insurance claims has potential for selection bias and less exact matching. These could lead to underestimation of effect sizes (uninsured persons with severe diseases like SCD that are highly represented in Medicaid would be expected to have higher complication rates than those with insurance) and overestimation of uncertainty (deidentification of Marketscan data limits the number of variables that are available for matching).[49] Efforts to replicate the findings of this study would benefit from utilizing data from a setting with universal health care and/or integrating data from both commercial claims and Centers for Medicare & Medicaid Services (CMS) beneficiaries. Third, our analysis is not able to account for differences in healthcare utilization overall or account for the impact of interventions such as hydroxyurea or newborn screening.[50] The meta-regression approach that we employed will accommodate future integration and synthesis of multiple data sources, which may continue to strengthen the comprehensiveness of evidence of SCD complications. Fourth, while we were able to partially compensate for small case counts for some outcomes with careful data processing and modelling approaches, there are a number of outcomes with plausible associations to SCD that we could not study because they are not routinely treated within the US health-care system and/or not documented using ICD codes, such as mental health comorbidities,[51,52] oral disorders, anthropometry and metabolic risk factors, and infectious diseases not commonly seen in the USA.[53–55] Our analysis of pregnancy outcomes further could not account for the potential differential fertility in people with SCD, which may have biased assessments of IRR for pregnancy complications.

The implications of the results extend well beyond the geographic setting that produced them. Namely, our study – the first to comprehensively quantify the health impacts of SCD as a risk factor by age and sex – highlights the massive gap in population health assessments on the impact of conditions like SCD. Global benchmarking frameworks such as the GBD Study consider “causes” and “risk factors” separately. For causal analyses of diseases and injuries, indirect burden is ignored and instead, burden is only counted when death and disability are directly and uniquely assigned to a given condition. This “one death one cause” paradigm can be a useful heuristic for accounting purposes when only limited data exist, but ends up fundamentally misrepresenting the impacts of severe systemic diseases like SCD where direct burden is only a small proportion of the total, as we have previously demonstrated[6].

While the indirect burden due to risk factors can be quantified via comparative-risk assessment (CRA) frameworks, GBD and similar efforts have historically restricted CRA to only a subset of risk factors historically considered ‘modifiable’ via population-level policy interventions.[56] In demonstrating the measurable, large, and significant excess risk of outcomes associated with exposure to SCD, our study provides a key building block to allow for full quantification of indirect burden and address the massive underestimation of SCD burden globally. We expect that full enumeration of SCD burden clearly demonstrates how a comprehensive SCD strategy is needed to achieve national and global public health targets such as those espoused in the Sustainable Development Goals.

SCD is a lifelong, multiorgan disease with numerous complications to screen starting at birth and continuing throughout the lifespan. Multidisciplinary approaches are required to ensure universal screening for SCD, close follow-up and evaluation for complications, and provision of appropriate preventive care and treatment. The magnitude of associated health risks should be concerning for health-care providers, health systems, and policymakers seeking to improve individual and population-level health outcomes.

## Data Availability

The data underlying this study are individual-level health insurance claims from the MarketScan Commercial Claims and Encounters and Medicare Supplemental and Coordination of Benefits databases (2016–2019). Merative owns these databases and licensed them to the authors. The license prohibits the authors from directly sharing the data, or any data derived from them at anything less than national level. The aggregated, unlabeled/ deidentified model inputs can be visualized in the Supporting Information file along with pooled model fits. Researchers can obtain the same data by licensing them directly from Merative the authors had no special access privileges. All estimates reported in this study appear in the tables and figures of the article (and the supplement). The analytic code used to generate the estimates will be made publicly available on the Global Health Data Exchange (https://ghdx.healthdata.org).

